# Cardiovascular Risk Stratification Among University Employees in India: Comparison of WHO/ISH, ASCVD and Framingham Risk Models

**DOI:** 10.64898/2026.10.12.26365229

**Authors:** Abhijeet Jagtap, Archana Ingole Patil, Abhay Kudale

## Abstract

**Background:** Cardiovascular disease (CVD) risk prediction is an important component of primary prevention, particularly in populations with increasing cardiometabolic risk. Several risk prediction models are available, but their estimates may differ because of differences in derivation populations, predictors, calibration, and cardiovascular outcomes. Evidence comparing these models within Indian occupational populations remains limited. This study compared 10-year CVD risk estimates generated by the World Health Organization/International Society of Hypertension (WHO/ISH) cardiovascular risk charts, the American College of Cardiology/American Heart Association (ACC/AHA) Pooled Cohort Equation (ASCVD-PCE), and the Framingham general cardiovascular disease risk equation among university employees in India.

**Methods:** A comparative observational study was conducted using standardized health assessment data from 266 university employees. Participants aged 40–74 years were selected to provide a common analytical age range across the three risk prediction approaches. Of 127 employees within this age range, three with documented previous ischemic heart disease were excluded because the analysis focused on primary prevention. The final analytical cohort comprised 124 participants. Blood pressure was assessed using a standardized protocol involving three readings at five-minute intervals, with repeat assessments during three subsequent weekly visits for employees with elevated initial readings. Ten-year cardiovascular risk was estimated using the 2019 WHO/ISH laboratory-based risk charts for the South Asian region, the ASCVD-PCE, and the 2008 Framingham general CVD equation. Agreement between risk categories was assessed using Cohen’s kappa and linear weighted kappa. Agreement was also examined at ≥10% and ≥20% thresholds.

**Results:** The mean age of the 124 participants was 47.63 ± 6.45 years, and 86.3% were male. Diabetes was present in 25.8%, smoking/tobacco use in 19.4%, and 29.0% were receiving antihypertensive treatment. Mean estimated risk was 5.94% for WHO/ISH, 5.95% for ASCVD-PCE, and 12.86% for Framingham. High risk (≥20%) was identified in 2.4%, 4.8%, and 20.2% of participants by WHO/ISH, ASCVD-PCE, and Framingham, respectively. Exact four-category agreement was 70.2% between WHO/ISH and ASCVD-PCE, compared with 31.5% between WHO/ISH and Framingham and 26.6% between ASCVD-PCE and Framingham. Cohen’s kappa was 0.502 for WHO/ISH versus ASCVD-PCE, 0.079 for WHO/ISH versus Framingham, and 0.047 for ASCVD-PCE versus Framingham. Weighted kappa values were 0.639, 0.323, and 0.342, respectively.

**Conclusion:** WHO/ISH and ASCVD-PCE produced closely similar mean risk estimates and substantially greater categorical agreement than either model showed with the Framingham equation. Framingham generated considerably higher 10-year CVD risk estimates and classified a larger proportion of participants into higher-risk categories. These findings highlight the importance of model selection when cardiovascular risk assessment is undertaken in Indian occupational populations and support cautious interpretation of risk estimates derived from models developed in different populations and for different cardiovascular endpoints.

## 1. Introduction

Cardiovascular diseases (CVDs) remain the leading cause of death globally and represent a substantial public health burden, particularly in low- and middle-income countries. The World Health Organization estimated that approximately 19.8 million people died from CVDs in 2022, accounting for about 32% of all global deaths, with more than three-quarters of CVD deaths occurring in low- and middle-income countries.[1] Most CVDs are attributable to modifiable behavioural and metabolic risk factors, including tobacco use, unhealthy diet, physical inactivity, harmful alcohol use, obesity, hypertension, diabetes, and dyslipidaemia.[1,2] Because these risk factors frequently cluster during the working years, early identification of individuals at elevated cardiovascular risk is an important component of preventive health programmes.

South Asian populations are of particular concern because of their substantial burden of cardiovascular risk factors and the occurrence of cardiovascular events at relatively younger ages. However, cardiovascular risk prediction remains challenging because many widely used risk equations were developed in populations that differ substantially from South Asian populations in baseline cardiovascular risk, risk-factor distributions, and event patterns.[3,4] A systematic review of cardiovascular risk models applicable to South Asian populations identified remarkably few studies formally evaluating model performance and concluded that further research was required to develop and validate risk models specifically for South Asian populations.[3] A subsequent review similarly highlighted uncertainty regarding the accuracy and transportability of commonly used cardiovascular risk calculators in South Asians and emphasized the need for large prospective South Asian cohorts.[4]

This concern is particularly relevant to India, where cardiovascular disease represents a major public health burden and where recalibration studies have demonstrated that predicted risk can vary substantially according to the population characteristics and baseline event rates used in the model.[5,6] For example, comparison of original and recalibrated Framingham models using Indian urban population data demonstrated substantial differences in the proportions classified as high risk, emphasizing the potential consequences of applying non-local prediction equations without recalibration.[5]

Risk prediction models provide an opportunity to integrate multiple cardiovascular risk factors into an estimate of an individual’s absolute risk over a defined period, most commonly 10 years. Such estimates can assist in identifying individuals who may benefit from intensified lifestyle modification, clinical assessment, pharmacological prevention, or closer follow-up. However, the clinical usefulness of a risk prediction model depends not only on its statistical performance but also on the population in which it is applied and the extent to which its predictions remain calibrated. [3-5]

Several cardiovascular risk prediction approaches are currently used internationally. The World Health Organization (WHO) developed updated cardiovascular disease risk prediction models and regional risk charts in 2019 to facilitate risk estimation across 21 global regions.[7] These models incorporate regional information and provide estimates for South Asia, making the WHO/ISH approach particularly relevant to countries such as India.

The American College of Cardiology/American Heart Association (ACC/AHA) Pooled Cohort Equation (PCE) was developed to estimate the 10-year risk of first hard atherosclerotic cardiovascular disease (ASCVD) events, including myocardial infarction, coronary heart disease death, and fatal or non-fatal stroke.[8] However, the PCE was derived primarily from US cohorts comprising non-Hispanic White and African American participants. Its application to South Asian populations may therefore involve important limitations. Evidence from a large integrated healthcare system has suggested that the PCE may underestimate cardiovascular risk among South Asian individuals, particularly among those classified at lower predicted risk.[9]

The Framingham general cardiovascular disease risk equation is another widely used prediction approach. The 2008 Framingham model estimates general CVD risk using age, total cholesterol, HDL cholesterol, systolic blood pressure, antihypertensive treatment, smoking, and diabetes.[10] Unlike the ASCVD-PCE, the Framingham general CVD model includes a broader range of cardiovascular outcomes, including coronary heart disease, cerebrovascular disease, peripheral arterial disease, and heart failure.

Although these models share several predictors, they differ in derivation populations, statistical methodology, endpoint definitions, and calibration. Consequently, the same individual may receive substantially different risk estimates depending on the model used. Previous South Asian research has demonstrated differences between Framingham and WHO/ISH classifications,[11] while studies from India have shown variation between original and recalibrated risk equations.[5,6] More recent evidence reinforces this concern. A 2026 study involving 4,975 South Asian patients presenting with a first acute myocardial infarction found substantial differences in risk classification across Framingham, ASCVD, WHO, JBS-3, and PREVENT models.[12] Although that population differed substantially from the occupational population studied here, the findings demonstrate that major cardiovascular risk algorithms may classify the same South Asian individuals quite differently.

The occupational setting provides an important opportunity for early identification of cardiovascular risk because employees may undergo periodic health assessment before development of clinically overt cardiovascular disease. University employees represent a relatively stable occupational population in which anthropometric, biochemical, behavioural, and clinical risk factors can be assessed systematically.

A recent Indian tertiary-care study comparing WHO/ISH, ASCVD, and Framingham risk scores demonstrated substantial differences in risk classification, particularly between Framingham and the other two approaches.[13] However, clinical NCD clinic populations may differ considerably from relatively healthy occupational populations. There remains limited evidence comparing these three risk prediction approaches within the same Indian occupational cohort using standardized clinical and biochemical measurements.

Therefore, the present study aimed to compare 10-year cardiovascular risk estimates generated by WHO/ISH, ASCVD-PCE, and Framingham models among university employees in India and to assess the extent of agreement between these approaches. The study specifically examined whether the three models classified the same individuals into comparable risk categories when applied to the same clinical and biochemical dataset.

## 2. Materials and Methods

### 2.1 Study design and setting

A comparative observational study was conducted using health assessment data of university employees attending the University Health Centre of Punyashlok Ahilyadevi Holkar Solapur University, Maharashtra, India. The study compared three established cardiovascular disease (CVD) risk prediction approaches—the World Health Organization/International Society of Hypertension (WHO/ISH) cardiovascular risk charts, the American College of Cardiology/American Heart Association (ACC/AHA) Pooled Cohort Equation (ASCVD-PCE), and the Framingham general cardiovascular disease risk equation for estimation of 10-year cardiovascular risk.

The analysis was based on standardized clinical, anthropometric, and biochemical measurements obtained as part of the institutional employee health assessment programme. The primary analysis was cross-sectional, with standardized repeat blood pressure assessments undertaken among employees with elevated blood pressure during initial screening.

### 2.2 Study population and eligibility

The source dataset comprised 266 university employees who underwent institutional health assessment. For comparative cardiovascular risk estimation, participants aged **40-74 years** were considered eligible. This age range was selected to permit application of all three risk prediction approaches within a common analytical population.

The 2019 WHO cardiovascular risk models are designed for cardiovascular risk estimation beginning at 40 years of age, while the ASCVD Pooled Cohort Equations were developed for adults aged 40-79 years. In contrast, the Framingham general cardiovascular disease risk equation was derived in participants aged 30-74 years. [7-10] Accordingly, 40-74 years represented the common applicable analytical age range for the three models.

Of the 266 employees in the source dataset, 127 were aged 40-74 years. Three participants with documented previous ischemic heart disease (IHD) were subsequently excluded because the present analysis focused on primary prevention risk estimation rather than risk prediction among individuals with established cardiovascular disease. The final analytical cohort therefore comprised 124 participants.

The sample size of 124 was determined by the number of eligible participants available within the predefined common age range after exclusion of participants with established IHD; it was not a minimum sample-size requirement for calculation of individual cardiovascular risk scores. All three risk prediction models were applied to the same 124 participants using the individual-level variables required by each model. This common-cohort approach enabled direct comparison of risk estimates and risk-category agreement between the three prediction approaches.

### 2.3 Health assessment and blood pressure measurement

During the institutional health assessment, trained healthcare personnel collected information on personal and medical history, cardiovascular risk factors, medication use, lifestyle characteristics, anthropometric measurements, blood pressure, and biochemical parameters.

Blood pressure was measured by trained nursing personnel using a validated automated blood pressure monitor (OMRON HBP-1300). Participants were instructed to avoid smoking, caffeine consumption, and strenuous physical activity before measurement. Following a 5-minute seated rest period, three blood pressure readings were obtained at 5-minute intervals, with the participant seated comfortably and the arm supported at heart level. The average of the three readings was used for the initial assessment.

Employees with systolic blood pressure (SBP) ≥140 mmHg and/or diastolic blood pressure (DBP) ≥90 mmHg during the initial screening were invited for repeat blood pressure assessment during three subsequent weekly visits. The same standardized measurement protocol was followed at each visit.

Participants whose blood pressure remained ≥140/90 mmHg during the follow-up assessments and who had no previous history of hypertension were classified as having newly detected hypertension. Employees with blood pressure values below this threshold and no previous diagnosis of hypertension were classified as normotensive.

Employees identified with elevated blood pressure were counselled and referred for appropriate medical evaluation and management. For cardiovascular risk estimation, the relevant systolic blood pressure measurement and antihypertensive treatment status required by each risk prediction model were used.

### 2.4 Anthropometric and biochemical assessment

Anthropometric assessment included body mass index (BMI) and waist-to-hip ratio (WHR). Biochemical assessment included fasting blood glucose, postprandial blood glucose, glycated haemoglobin (HbA1c), total cholesterol, triglycerides, high-density lipoprotein cholesterol (HDL-C), and low-density lipoprotein cholesterol (LDL-C), as available in the institutional health assessment dataset. Diabetes was considered present when there was a documented history of diabetes and/or HbA1c ≥6.5%. Current antihypertensive and statin treatment status was determined from the documented medication history.

### 2.5 Cardiovascular risk prediction models

### 2.5.1 WHO/ISH cardiovascular risk charts

The 2019 WHO cardiovascular disease risk prediction charts were used with the laboratory-based model applicable to the South Asian region.[7] The WHO models estimate the 10-year probability of a fatal or non-fatal cardiovascular event, including myocardial infarction and stroke.

The laboratory-based model incorporates age, sex, smoking status, systolic blood pressure, diabetes status, and total cholesterol. Risk estimates were categorized as &[lt]5%, 5-&[lt]10%, 10-&[lt]20%, and ≥20%. For comparative analysis, the original WHO categories of 20-&[lt]30% and ≥30% were combined into a single ≥20% category.

#### 2.5.2 ASCVD Pooled Cohort Equation

The 2013 ACC/AHA Pooled Cohort Equation was used to estimate 10-year risk of a first hard atherosclerotic cardiovascular disease (ASCVD) event.[8] The Pooled Cohort Equations were developed for adults aged 40-79 years and incorporate age, sex, total cholesterol, HDL-C, systolic blood pressure, antihypertensive treatment, smoking, and diabetes status.

The ASCVD endpoint includes non-fatal myocardial infarction, coronary heart disease death, and fatal or non-fatal stroke. Because the original Pooled Cohort Equations were developed using non-Hispanic White and African American populations and no India-specific PCE is available, the sex-specific White equation was used as a standardized computational approach for the Indian participants. The ACC/AHA guideline allows consideration of the White equation when estimating risk in populations other than African American and White populations.[8] This application was treated as a comparative risk-estimation approach and does not imply biological equivalence between the Indian study population and the White derivation population.

#### 2.5.3 Framingham general cardiovascular disease risk equation

The 2008 Framingham general cardiovascular disease risk equation was used to estimate 10-year general CVD risk.[10] The original equation was derived among participants aged 30-74 years who were free of cardiovascular disease at baseline. The model incorporates age, total cholesterol, HDL-C, systolic blood pressure, antihypertensive treatment, smoking, and diabetes. Its general CVD endpoint includes coronary heart disease, cerebrovascular disease, peripheral arterial disease, and heart failure. The Framingham equation was applied without population-specific recalibration.

### 2.6 Rationale for the common analytical age range

The three risk prediction approaches differ in their original derivation populations, endpoints, and age ranges. The WHO 2019 models include adults beginning at 40 years, the ASCVD-PCE was developed for adults aged 40-79 years, and the Framingham general CVD equation was derived for individuals aged 30-74 years. [7-10] Therefore, the study restricted the comparative analysis to 40-74 years, representing the common age range in which all three approaches could be applied. Importantly, this restriction was implemented to ensure methodological comparability across models and not because a sample size of 124 participants was required for cardiovascular risk calculation. Individual cardiovascular risk can be estimated for an eligible person independently of the eventual number of participants in a research study. The final sample of 124 represented the complete eligible analytical cohort available from the institutional dataset after application of the prespecified age and disease-status criteria.

### 2.7 Harmonization of risk categories

To permit direct comparison between the three prediction approaches, individual risk estimates were classified into four common categories: &[lt]5%, 5-&[lt]10%, 10-&[lt]20% and ≥20%. These categories were used consistently for cross-classification and agreement analyses. Because the three models predict different cardiovascular endpoints, their estimates were interpreted as 10-year estimated cardiovascular risk rather than as directly interchangeable measures of an identical outcome. The analysis therefore assessed agreement and concordance between models, rather than determining which model was more accurate. In the absence of prospective cardiovascular outcome follow-up, differences in risk estimates were not interpreted as evidence of model overestimation or underestimation.

### 2.8 Statistical analysis

Continuous variables were summarized using mean and standard deviation (SD), while categorical variables were summarized using frequencies and percentages. The distribution of 10-year risk estimates generated by each model was summarized using mean, SD, median, and range. Cross-tabulations were constructed to compare risk-category classifications between pairs of models. Cohen’s kappa (κ) was used to assess agreement between models based on the four-category risk classification. Because the risk categories were ordinal, linear weighted kappa was additionally calculated to account for the magnitude of disagreement between categories. Agreement was further examined using dichotomous risk thresholds of ≥10% and ≥20%. Percentage agreement and Cohen’s kappa were calculated for each threshold. Paired comparisons of continuous risk estimates between models were performed using the paired t-test and the Wilcoxon signed-rank test. Two-sided *p* values &[lt]0.05 were considered statistically significant. Sensitivity analyses were conducted after excluding participants with diabetes, participants receiving antihypertensive treatment, participants with both diabetes and treated hypertension, and smokers, to examine the robustness of agreement between the risk prediction models. All three models were applied to the identical analytical cohort of 124 participants, thereby minimizing differences arising from variation in the underlying study population when comparing model-derived estimates.

### 2.9 Ethical considerations

Ethical approval was obtained from the Institutional Ethics Committee of Savitribai Phule Pune University ((Approval No. SPPU/IEC/2019/07).

## 3. Results

### 3.1 Participant selection and baseline characteristics

Of the 266 university employees included in the institutional health assessment dataset, 127 were aged 40-74 years. Three participants with documented previous IHD were excluded from the primary prevention analysis, resulting in a final analytical cohort of 124 participants. (Ref. Fig.1) The mean age was 47.63 ± 6.45 years, with a range of 40-70 years. The majority of participants were male (107; 86.3%). Mean SBP was 135.09 ± 17.98 mmHg, while mean DBP was 83.61 ± 11.95 mmHg. Mean BMI was 25.76 ± 3.83 kg/m^2^. Over half of participants (54.0%) were classified as overweight and a further 8.9% as obese. Diabetes was present in 32 participants (25.8%), while 24 (19.4%) reported smoking or tobacco use and 26 (21.0%) reported alcohol use. Thirty-six participants (29.0%) were receiving antihypertensive treatment. Mean total cholesterol was 192.29 ± 39.25 mg/dL, while mean HDL-C was 43.93 ± 11.97 mg/dL and mean LDL-C was 130.60 ± 38.80 mg/dL. The baseline characteristics of the 124 participants included in the analysis are presented in Table 1

**Table 1.** Baseline characteristics of the analytical cohort (n=124)

| Characteristic | Value |
| --- | --- |
| Age, years, mean ± SD | $47.63 \pm 6.45$ |
| Male, n (%) | 107 (86.3) |
| Female, n (%) | 17 (13.7) |
| SBP, mmHg, mean ± SD | $135.09 \pm 17.98$ |
| DBP, mmHg, mean ± SD | $83.61 \pm 11.95$ |
| BMI, kg/m<sup>2</sup>, mean ± SD | $25.76 \pm 3.83$ |
| Underweight, n (%) | 2 (1.6) |
| Normal BMI, n (%) | 44 (35.5) |
| Overweight, n (%) | 67 (54.0) |
| Obese, n (%) | 11 (8.9) |
| Diabetes, n (%) | 32 (25.8) |
| Smoking/tobacco use, n (%) | 24 (19.4) |
| Alcohol use, n (%) | 26 (21.0) |
| Current antihypertensive treatment, n (%) | 36 (29.0) |
| Current statin treatment, n (%) | 36 (29.0) |
| Total cholesterol, mg/dL, mean ± SD | $192.29 \pm 39.25$ |
| HDL-C, mg/dL, mean ± SD | $43.93 \pm 11.97$ |
| LDL-C, mg/dL, mean ± SD | $130.60 \pm 38.80$ |
| Waist-to-hip ratio, mean ± SD | $0.948 \pm 0.031$ |
| Perceived stress score, mean ± SD | $15.97 \pm 5.95$ |
| Sitting time, hours/day, mean ± SD | $5.27 \pm 2.36$ |
| Service experience, years, mean ± SD | $16.78 \pm 9.16$ |

(SD, standard deviation; SBP, systolic blood pressure; DBP, diastolic blood pressure; BMI, body mass index; HDL-C, high-density lipoprotein cholesterol; LDL-C, low-density lipoprotein cholesterol.)

**Fig. 1.**
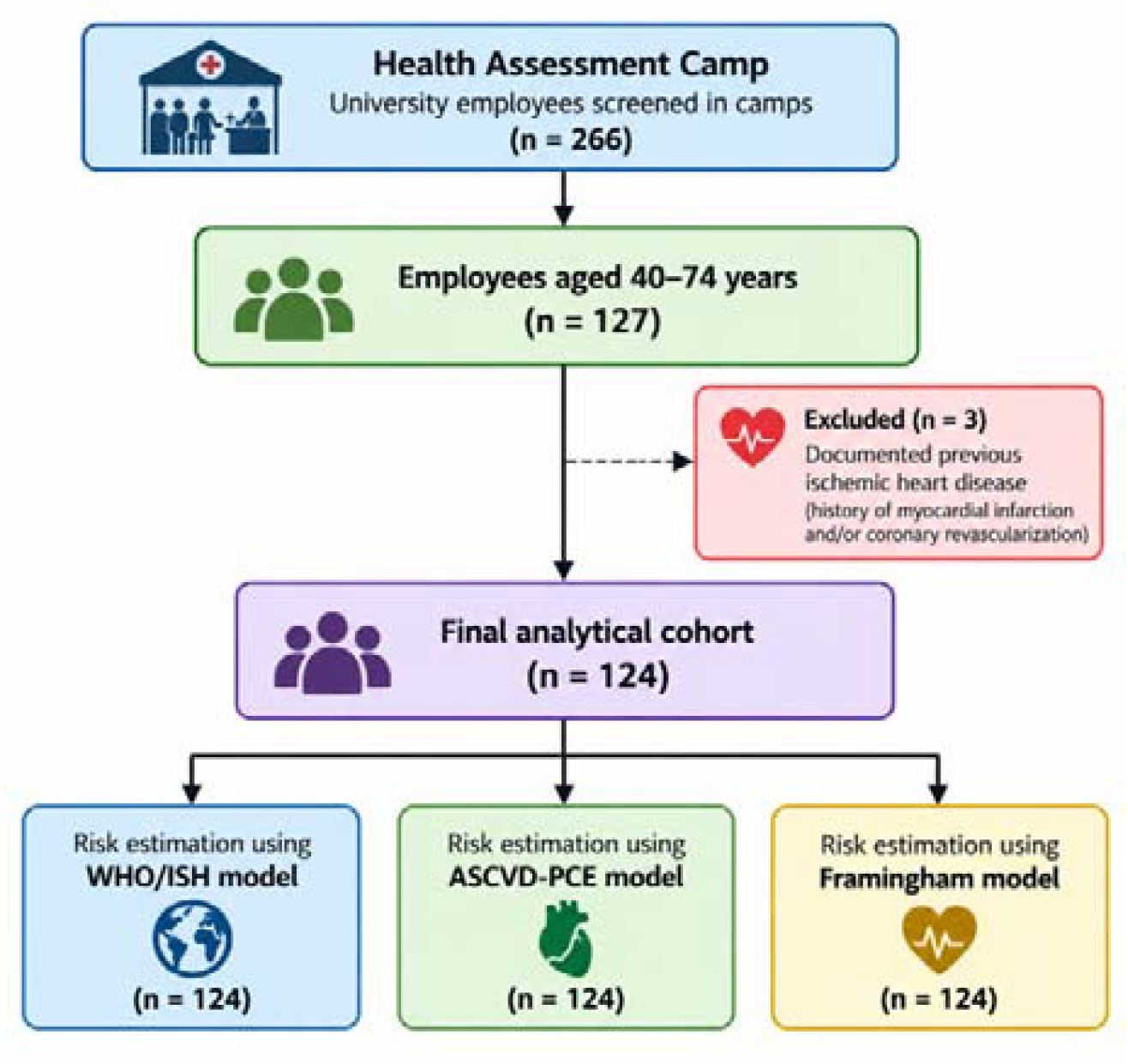
Participant Selection and Analytical Pathway.

### 3.2 Distribution of cardiovascular risk estimates

The distribution of participants across the four harmonized 10-year cardiovascular risk categories differed substantially between the three models (Table 2; Figure 2).

**Table 2.**
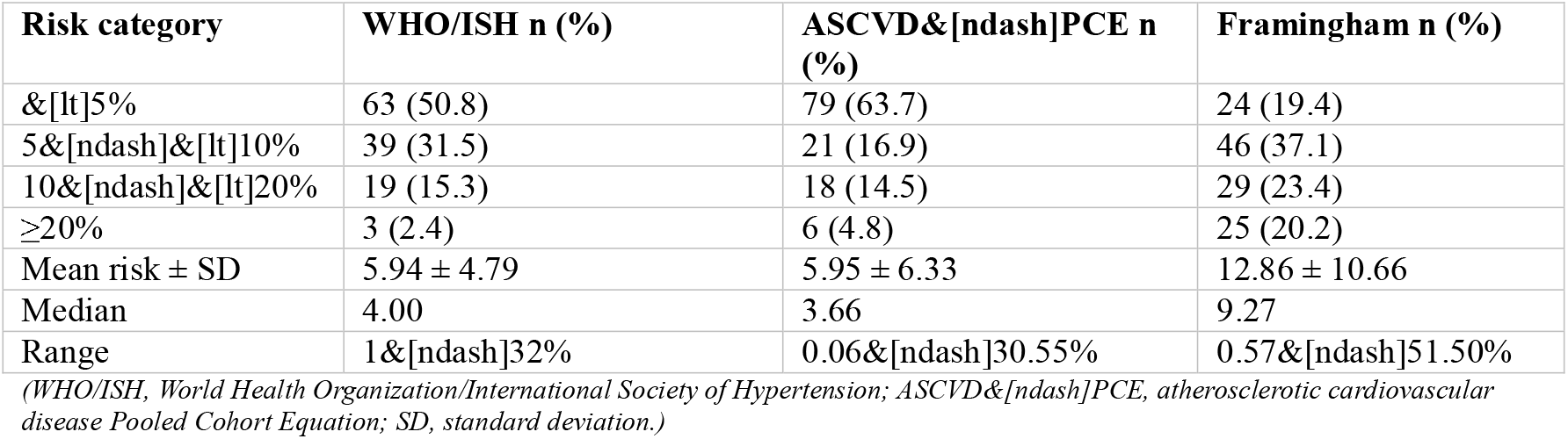
Distribution of 10-year cardiovascular risk estimates according to prediction model.

**Table 3.**
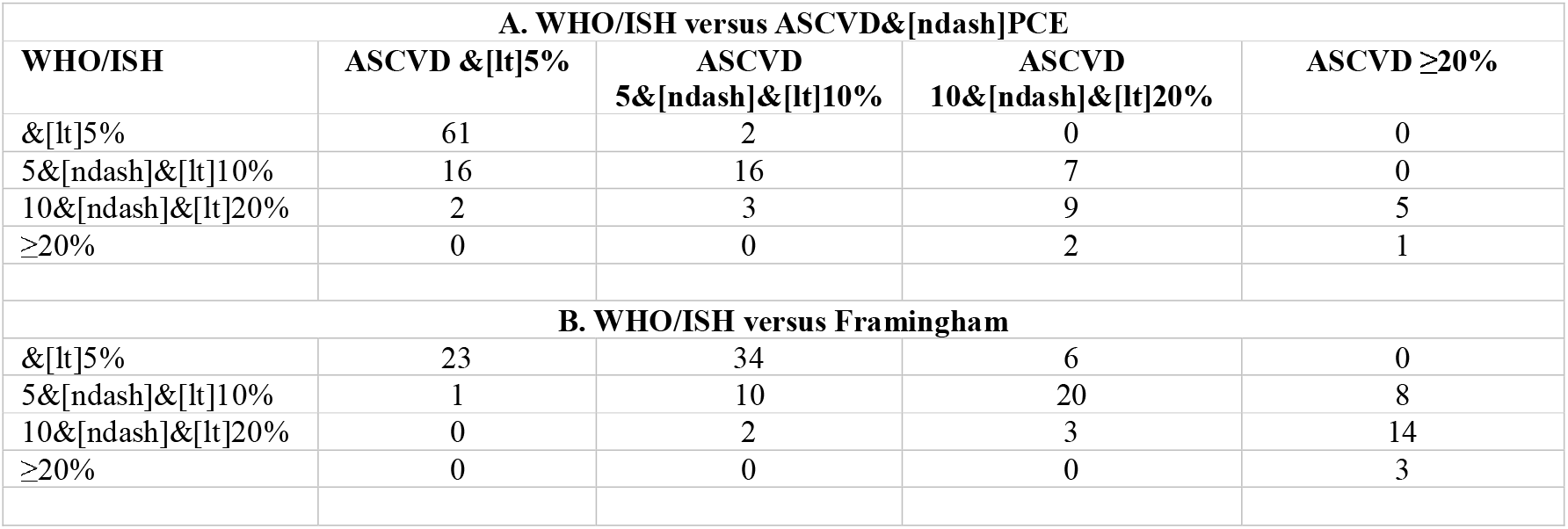
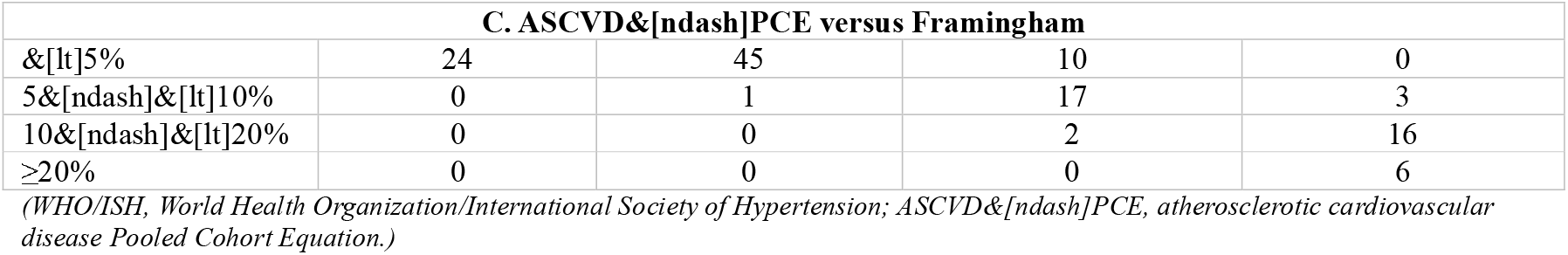
Cross-classification of participants according to pairs of cardiovascular risk prediction models.

**Table 4.** Agreement between cardiovascular risk prediction models.

| Comparison | Exact agreement, % | Cohen's $\kappa$ | Linear weighted $\kappa$ |
| --- | --- | --- | --- |
| WHO/ISH vs ASCVD&[ndash]PCE | 70.16 | 0.502 | 0.639 |
| WHO/ISH vs Framingham | 31.45 | 0.079 | 0.323 |
| ASCVD&[ndash]PCE vs Framingham | 26.61 | 0.047 | 0.342 |

**Table 5.** Sensitivity analyses of agreement between cardiovascular risk prediction models.

| Analysis | n | WHO/ISH vs<br>ASCVD&[ndash]P<br>CE $\kappa$ | Weighte<br>d $\kappa$ | WHO/IS<br>H vs FRS<br>$\kappa$ | Weighte<br>d $\kappa$ | ASCVD&[ndash]P<br>CE vs FRS $\kappa$ | Weighte<br>d $\kappa$ |
| --- | --- | --- | --- | --- | --- | --- | --- |
| All participants | 124 | 0.502 | 0.639 | 0.079 | 0.323 | 0.047 | 0.342 |
| No diabetes | 92 | 0.629 | 0.713 | 0.017 | 0.222 | 0.020 | 0.263 |
| No treated hypertension | 88 | 0.473 | 0.605 | 0.169 | 0.354 | 0.045 | 0.304 |
| No diabetes + no treated hypertension | 75 | 0.600 | 0.684 | 0.056 | 0.242 | 0.032 | 0.268 |
| Non&[ndash]smokers | 100 | 0.517 | 0.637 | 0.086 | 0.298 | 0.055 | 0.294 |

**Table 6.**
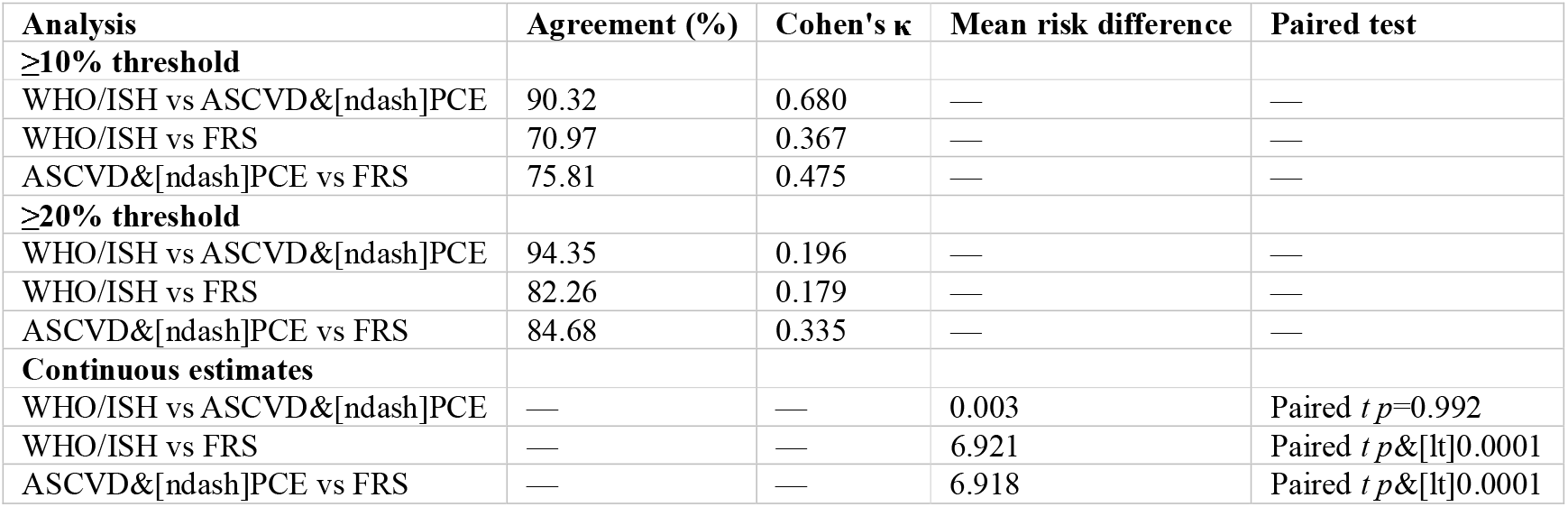
Threshold-based agreement and paired comparisons of continuous risk estimates.

**Fig. 2.**
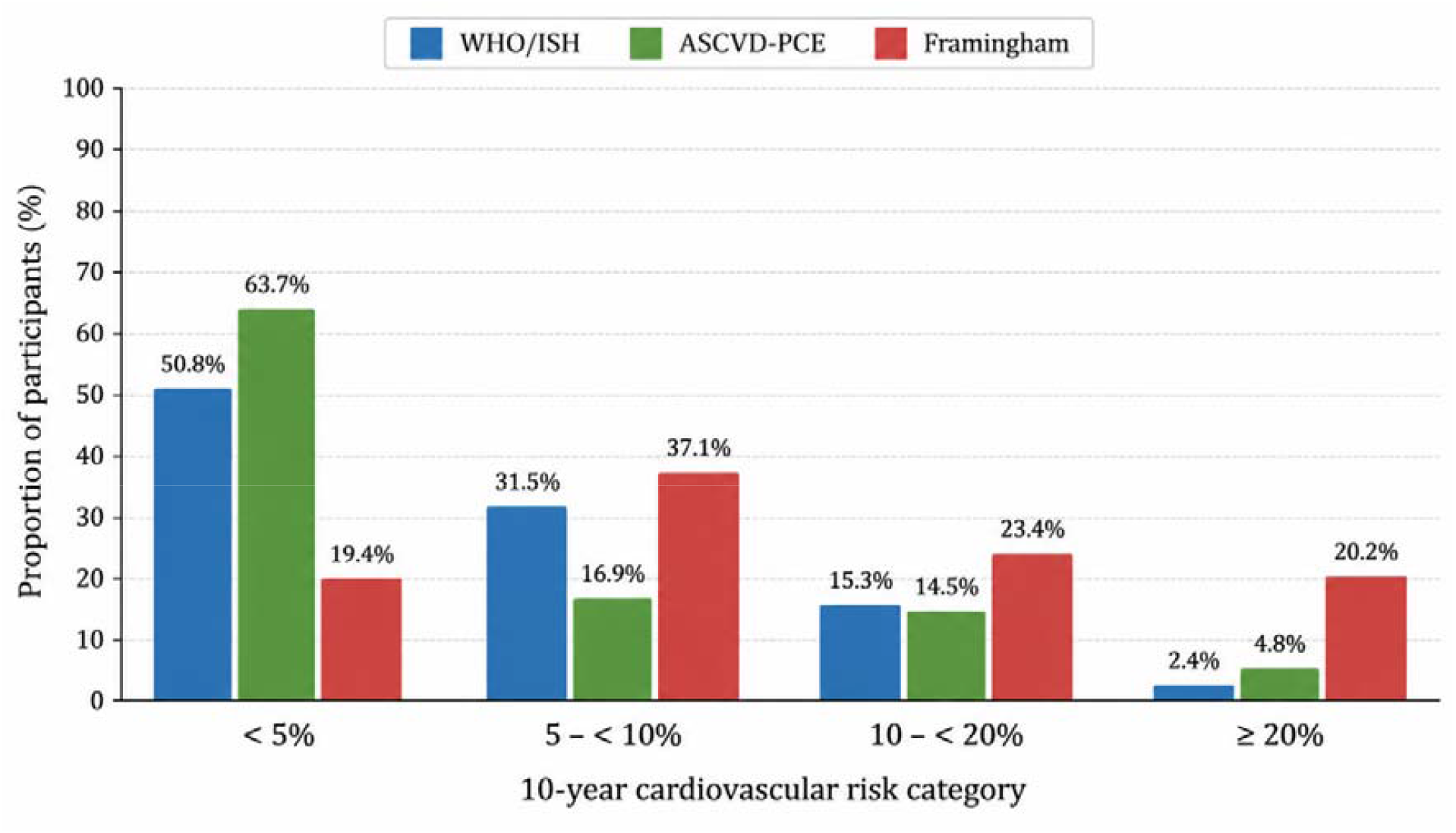
Distribution of Participants across 10-year cardiovascular risk categories according to WHO/ISH, ASCVD-PCE and Framingham Models.

Using WHO/ISH, 63 participants (50.8%) were classified as having a predicted risk &[lt]5%, 39 (31.5%) as 5-&[lt]10%, 19 (15.3%) as 10-&[lt]20%, and 3 (2.4%) as ≥20%. The ASCVD-PCE classified 79 participants (63.7%) as &[lt]5%, 21 (16.9%) as 5-&[lt]10%, 18 (14.5%) as 10-&[lt]20%, and 6 (4.8%) as ≥20%. In contrast, the Framingham equation classified only 24 participants (19.4%) as &[lt]5%, while 46 (37.1%) were classified as 5-&[lt]10%, 29 (23.4%) as 10-&[lt]20%, and 25 (20.2%) as ≥20%. The mean risk estimates were 5.94% for WHO/ISH, 5.95% for ASCVD-PCE, and 12.86% for Framingham. The median values were 4.00%, 3.66%, and 9.27%, respectively. (Ref. Table 2) The WHO/ISH and ASCVD-PCE mean estimates were almost identical, with a mean difference of only 0.003 percentage points. In contrast, the Framingham estimate was approximately 6.9 percentage points higher than either WHO/ISH or ASCVD-PCE.

### 3.3 Cross-classification between risk prediction models

The cross-classification matrices demonstrated substantial differences in individual risk categorization. Between WHO/ISH and ASCVD-PCE, 61 participants were classified as &[lt]5% by both models, while 16 were classified as 5-&[lt]10% by both. Nine participants were classified as 10-&[lt]20% by both models. Overall, 87 of the 124 participants were placed in the same risk category by the two approaches.

For WHO/ISH versus Framingham, exact agreement was considerably lower. Only 23 participants were classified as &[lt]5% by both models, while 34 WHO/ISH participants classified as &[lt]5% were classified as 5-&[lt]10% by Framingham.

Similarly, ASCVD-PCE and Framingham showed relatively limited exact agreement, with most disagreement occurring because Framingham assigned participants to higher risk categories.

### 3.4 Agreement between cardiovascular risk prediction models

The greatest agreement was observed between WHO/ISH and ASCVD-PCE. Exact four-category agreement was 70.2%, with Cohen’s κ of 0.502 and linear weighted κ of 0.639. Agreement between WHO/ISH and Framingham was substantially lower, with exact agreement of 31.5%, Cohen’s κ of 0.079, and weighted κ of 0.323. ASCVD-PCE and Framingham showed the lowest exact agreement at 26.6%, with Cohen’s κ of 0.047 and weighted κ of 0.342.

### 3.5 Sensitivity analyses

The greater agreement between WHO/ISH and ASCVD-PCE remained evident across sensitivity analyses. Among participants without diabetes (n=92), Cohen’s κ increased to **0.629** for WHO/ISH versus ASCVD-PCE, with weighted κ of 0.713. Among participants without treated hypertension (n=88), the corresponding values were 0.473 and 0.605. Among participants without both diabetes and treated hypertension (n=75), WHO/ISH versus ASCVD-PCE showed Cohen’s κ of 0.600 and weighted κ of 0.684. Among non-smokers (n=100), Cohen’s κ was 0.517 and weighted κ was 0.637. The agreement between WHO/ISH and Framingham and between ASCVD-PCE and Framingham remained low across these sensitivity analyses.

### 3.6 Threshold-based agreement and paired comparison of continuous estimates

When the clinically relevant threshold of ≥**10%** was applied, agreement improved across all three model comparisons. WHO/ISH versus ASCVD-PCE showed 90.3% agreement and κ=0.680. WHO/ISH versus Framingham showed 71.0% agreement and κ=0.367, while ASCVD-PCE versus Framingham showed 75.8% agreement and κ=0.475. At the ≥**20%** threshold, percentage agreement was high for all comparisons, although kappa values remained more modest because of the low prevalence of participants above this threshold in some models.

Paired comparison of continuous estimates showed no meaningful difference between WHO/ISH and ASCVD-PCE. The mean risk estimates were 5.9435% and 5.9464%, respectively, with a mean paired difference of only 0.0028 percentage points. The paired *t&*[ndash]test (*p*=0.992) and Wilcoxon signed-rank test (*p*=0.055) both indicated no statistically significant difference.

In contrast, Framingham generated significantly higher estimates than both WHO/ISH and ASCVD-PCE. The mean difference was approximately 6.92 percentage points in both comparisons, with both parametric and non-parametric paired tests yielding *p&*[lt]0.0001.

## 4. Discussion

This study compared three established cardiovascular risk prediction approaches within the same cohort of university employees in India. The principal finding was that WHO/ISH and ASCVD-PCE demonstrated substantially greater agreement with each other than either model demonstrated with the Framingham general CVD equation. Although the mean estimates generated by WHO/ISH and ASCVD-PCE were almost identical, Framingham produced substantially higher estimates and classified a considerably larger proportion of participants into higher-risk categories.

The close numerical similarity between WHO/ISH and ASCVD-PCE was notable. Their mean estimated risks were 5.94% and 5.95%, respectively, and the mean paired difference was essentially zero. Their four-category exact agreement was 70.2%, with a Cohen’s κ of 0.502 and weighted κ of 0.639. This indicates moderate unweighted agreement and stronger agreement when the ordinal nature of the risk categories is considered.

The high agreement between WHO/ISH and ASCVD-PCE is particularly relevant because the two models were developed using different approaches and for somewhat different endpoints. WHO/ISH provides a region-specific model intended to estimate the probability of fatal or non-fatal cardiovascular events, whereas ASCVD-PCE focuses on first hard ASCVD events. Thus, the observed similarity in this occupational cohort should not be interpreted as evidence that the models are mathematically or clinically identical. Rather, it indicates that they produced broadly comparable rankings and risk categories in this particular population.

In contrast, the Framingham equation generated considerably higher risk estimates. The mean Framingham estimate of 12.86% was more than twice the mean WHO/ISH and ASCVD-PCE estimates. Furthermore, 20.2% of participants were classified as having ≥20% risk by Framingham compared with 2.4% using WHO/ISH and 4.8% using ASCVD-PCE.

This finding is consistent with evidence from South Asian populations demonstrating differences between Framingham and WHO/ISH risk classifications.[11] In addition, Indian studies comparing original and recalibrated Framingham models have shown that the predicted proportion of individuals considered high risk can vary substantially depending on whether local risk-factor distributions and mortality data are incorporated.[5,6] These findings support the interpretation that differences between models may reflect calibration and transportability rather than simply differences in the mathematical computation of risk.

The differences between models may also be explained by their different outcome definitions. The Framingham general CVD model estimates a broader composite outcome that includes coronary heart disease, cerebrovascular disease, peripheral arterial disease, and heart failure.[10] The ASCVD-PCE estimates first hard ASCVD events, while WHO/ISH estimates fatal or non-fatal cardiovascular events using a region-specific model.[7,8] Consequently, the numerical estimates should not be regarded as interchangeable measures of exactly the same clinical endpoint.

The derivation populations also differ substantially. The WHO models were developed using multinational datasets and incorporate regional cardiovascular risk, whereas the PCE was derived primarily from US populations and the Framingham equation originated from the Framingham Heart Study cohort.[7,8,10] A systematic review of cardiovascular risk models applicable to South Asian populations found very limited evidence regarding formal model performance and emphasized the need for models developed and validated specifically in South Asian populations.[3] A subsequent review similarly highlighted uncertainty regarding the transportability of existing risk calculators to South Asians and emphasized the need for large prospective databases.[4]

This issue is particularly relevant to India. Gupta et al. demonstrated that recalibration of Framingham models using Indian urban population characteristics produced substantially different risk classifications compared with the original model.[5] Earlier work from rural India likewise demonstrated differences between the original Framingham equation and recalibrated models using local or national mortality and risk-factor data.[6] These findings indicate that applying an equation developed in another population without recalibration may produce materially different estimates of absolute cardiovascular risk.

The application of ASCVD-PCE in South Asians presents a similar concern. Mantri et al. reported evidence that the Pooled Cohort Equation underestimated risk among South Asian individuals, particularly among those classified at lower predicted risk.[9] More recently, Gupta et al. compared five cardiovascular risk prediction models among 4,975 South Asian patients presenting with first acute myocardial infarction and demonstrated substantial differences in pre-event risk classification across Framingham, ASCVD 2013, WHO, JBS-3, and PREVENT models.[12] Although that study involved patients with acute myocardial infarction and therefore differs substantially from the present primary-prevention occupational cohort, its findings independently reinforce the broader concern that contemporary cardiovascular risk algorithms can classify South Asian individuals quite differently.

The present study therefore does not interpret the similarity between WHO/ISH and PCE as evidence that the PCE is fully calibrated for Indian populations. Rather, the PCE was included as an internationally recognized comparator and applied using the standardized White equation because an India-specific PCE was unavailable.

The sensitivity analyses strengthened the principal finding. After excluding individuals with diabetes, WHO/ISH versus ASCVD-PCE agreement increased from κ=0.502 to 0.629. Similarly, among individuals without diabetes and treated hypertension, κ remained 0.600. The consistent pattern across subgroup analyses suggests that the relatively strong agreement between WHO/ISH and ASCVD-PCE was not driven exclusively by the presence of major individual risk factors.

The low agreement between Framingham and the other models persisted across sensitivity analyses. This suggests that the higher risk estimates produced by Framingham were not explained solely by diabetes, treated hypertension, or smoking.

Threshold-based analyses provided an additional perspective. At the ≥10% threshold, agreement between WHO/ISH and ASCVD-PCE increased to 90.3%, with κ=0.680. This indicates that despite some disagreement in the finer four-category classification, the two models frequently agreed when participants were dichotomized around a clinically relevant risk threshold.

However, the ≥20% threshold produced high percentage agreement but comparatively lower kappa values, particularly for WHO/ISH versus ASCVD-PCE. This illustrates an important statistical issue: when very few participants fall into a high-risk category, percentage agreement can be high even when chance-corrected agreement is modest. Therefore, both percentage agreement and kappa should be considered when interpreting categorical risk classification.

### 4.1 Occupational health implications

The findings have practical implications for cardiovascular risk screening in occupational health settings. University employees may appear relatively healthy because they are generally employed and remain within a working-age population; however, the present cohort demonstrated a substantial burden of cardiometabolic risk factors. These findings support the value of systematic cardiovascular risk assessment within institutional health programmes. Importantly, the choice of risk model substantially influenced risk classification. Framingham classified 20.2% of participants as having ≥20% 10-year risk, compared with only 2.4% with WHO/ISH and 4.8% with ASCVD-PCE. Such differences could affect counselling, referral, preventive treatment, and resource allocation. However, these findings do not establish the superiority of any model. In the absence of prospective cardiovascular outcomes, differences should be interpreted as variation in model-based risk estimates rather than differences in true cardiovascular risk.

### 4.2 Strengths

The study is strengthened by several methodological and contextual features. First, all three models were applied to the same 124 individuals, using the same underlying clinical and biochemical dataset. This eliminates a major source of variation that can occur when comparing risk scores across separate populations. Second, the study was conducted in an occupational health setting, which represents an important but relatively understudied setting for cardiovascular risk assessment. Third, blood pressure assessment followed a standardized protocol with three readings during initial screening and repeat assessment during three subsequent weekly visits for individuals with elevated blood pressure. This reduces reliance on a single elevated BP measurement. Fourth, the study evaluated both categorical agreement and continuous risk estimates. The use of weighted kappa was particularly useful because disagreement between adjacent risk categories is clinically different from disagreement across several categories. Finally, sensitivity analyses demonstrated that the principal pattern of agreement remained broadly consistent after exclusion of major cardiovascular risk-factor subgroups.

### 4.3 Limitations

This study has several limitations. First, the modest sample size (n=124), although representing the complete eligible institutional cohort, limits precision and generalizability. Second, the single-university setting and predominantly male sample may limit applicability to female employees and other occupational populations. Third, the cross-sectional design and absence of cardiovascular outcome follow-up precluded assessment of discrimination, calibration, or predictive accuracy; the study therefore evaluated agreement between predicted risks. Fourth, ASCVD-PCE was not derived or calibrated for Indian populations, and the use of the White equation was a standardized computational choice rather than an assumption of equivalent baseline risk. Fifth, Framingham estimates were not population-recalibrated; therefore, its higher estimates should be interpreted as model-generated values rather than definitive evidence of excess absolute risk. Sixth, the models predict somewhat different cardiovascular endpoints, limiting direct comparison of numerical estimates. Finally, despite repeated BP assessment, the observational design precludes causal inference.

## 5. Conclusion

Among Indian university employees aged 40-74 years, WHO/ISH and ASCVD-PCE produced closely similar 10-year cardiovascular risk estimates and showed substantially greater categorical agreement with each other than with Framingham. Framingham generated higher estimates and classified more participants into higher-risk categories. These differences likely reflect variations in derivation populations, endpoints, predictors, and calibration. The findings highlight that model selection can materially influence cardiovascular risk classification in Indian occupational populations. Larger prospective studies with representative cohorts and cardiovascular outcomes are needed to assess model discrimination, calibration, and clinical utility and to support India-specific or recalibrated risk equations.

## Data Availability

All data produced in the present study are available upon reasonable request to the authors

## Declarations

### Consent for publication

Not applicable, provided that the dataset contains no individually identifiable information.

### Availability of data and materials

The datasets generated and/or analyzed during the current study are not publicly available because they contain institutional employee health information but may be available from the corresponding author or institutional authority on reasonable request, subject to applicable ethical and institutional restrictions.

### Authors’ contributions

AJ, AI and AK conceptualized the study, contributed to study design, data analysis and interpretation, and drafted the manuscript. All authors contributed to interpretation of the findings, critically reviewed the manuscript, approved the final version, and agreed to be accountable for all aspects of the work.

### Declaration on use of Generative AI

During the preparation of this manuscript, the authors used a generative artificial intelligence (AI) tool (ChatGPT, OpenAI) to assist with language refinement, editing, organization of text, and improvement of readability. AI assistance was not used to generate, alter, or interpret the study data, perform statistical analyses, or make scientific or clinical decisions. All analyses, interpretations, and conclusions were independently reviewed and verified by the authors, who take full responsibility for the accuracy, integrity, and originality of the content. The authors also reviewed the manuscript for potential errors or inappropriate AI-generated content and approved the final version for submission.

